# Antibody Profiling and Prevalence in the US population during the SARS-CoV2 Pandemic

**DOI:** 10.1101/2020.04.29.20085068

**Authors:** Hari Krishnan Krishnamurthy, Vasanth Jayaraman, Karthik Krishna, Karenah E. Rajasekaran, Tianhao Wang, Kang Bei, John J. Rajasekaran, Inna Yaskin, Alex J. Rai, Rok Seon Choung, Joseph A. Murray

## Abstract

**Background:** Antibody diagnostics play an important role in disease detection and can potentially aid in monitoring of the immune responses to see if an individual has developed immunity. Developing high throughput diagnostics which does not involve handling of infectious material becomes imperative in the case of pandemics such as the recent outbreak of SARS-CoV2.

**Methods:** A protein microarray technology was used to detect the plurality of antibody response to four novel antigens namely S1 glycoprotein, Receptor binding domain (RBD), S2 glycoprotein and Nucleoprotein of the novel coronavirus named SARS-CoV2 using serum samples. A DBS card was additionally used to compare its performance with a venipuncture-based serum separator tube (SST) draw.

**Results:** The three main subclasses of antibodies IgM, IgA and IgG were analyzed to see the variations in immune responses in the affected population and compared to their microbial RT-PCR based NP swab results. The clinical sensitivity and specificity were determined to be 98.1% and 98.6%. In the matrix comparison study, which would enable patients to test without risk of transmitting the virus, DBS matched with higher than 98% accuracy to a venipuncture-based SST collection.

**Conclusion:** Multiplex testing enables higher sensitivity and specificity which is essential while establishing exposure on a population scale. This flexible platform along with a discrete collection methodology would be crucial and broadly useful to scale up testing in current and future pandemics. Minimum sample volume that can be collected using DBS cards can be processed in this multiplex pillar plate format enabling the capacity to provide the reliability of high throughput analyzers while having the ease of collection similar to rapid tests.

## INTRODUCTION

Early in December 2019, the first pneumonia cases of unknown origin were reported in Wuhan, China [1,2]. Subsequently this public health crisis that spread around the world has been identified to be caused by a novel coronavirus(2019-nCoV) or severe acute respiratory syndrome coronavirus 2(SARS-CoV-2) [3,4]. In most individuals that disease should resolve in itself but there are subgroups with higher risk of morbidity and mortality [5,6]. In severe cases massive alveolar damage and progressive respiratory failure has been reported [7]. In comparison to the previous coronavirus outbreaks the novel coronavirus appears to have higher transmissibility while having a somewhat higher mortality rate than seasonal flu making for a very toxic combination [8]. Additionally, limited availability of PCR testing for viral illness and the likelihood of a significant proportion of infected individuals having no or trivial symptoms might accelerate disease spread [9]

Diagnostic testing is paramount in identification, containment and monitoring of infectious disease outbreaks [10]. Ideally, the collection process should be performed in quarantine while the diagnostic assay should be highly automated with throughput sufficient to handle several thousands if not millions of samples per day. Antibody diagnostics have been a preferred and accurate method in detection and monitoring of many infectious diseases. Most antibody tests rely on traditional ELISA methodology which has limited multiplexing capability and excessive antigen requirement which limits their ability to be used at scale during pandemics [10]. Traditional methods of specimen collection from an infected individual increases the risk of disease spread and reduces testing capability due to limited availability of specimen collection personnel during such pandemics. We present here a flexible platform for multiplex detection of antibodies from affected individuals enabling high sensitivity with high specificity of detection. We also show comparison of traditional serum separator tube (SST) collection by phlebotomist with dried blood spot-based self-collection by affected subjects to provide for enhanced diagnostics availability during current and future pandemics

## MATERIALS AND METHODS

### Patient Population

Study 1. To obtain the antibody profile of individuals the SARS-CoV2 samples with swab based microbiological confirmation were collected from multiple healthcare centers. 1. Gunnison Valley hospital (# of samples: 132), 2. Elite Medical Center (# of samples: 26). In addition, a cohort of samples that were collected prior to the outbreak were used as negative controls along with disease controls. The sample cohort information age, gender and sample type collected are summarized in Table 1.

Study 2. Traditional phlebotomy-based SST collection comparison to Dried Blood spot collection by subject was performed at healthcare centers across the US. A cohort of 158 clinically paired samples were used for clinical comparison along with 1418 samples which were prospectively collected across the geographical locations in the US to identify infection spread and also validate dried blood spot collection vs venipuncture-based SST collection.

**Table 1:** Sample Cohort

| **Sample Type** | **N** | **Age, mean (Range)** | **Male %** | **Female %** |
| --- | --- | --- | --- | --- |
| **SARS-CoV-2 positive** | 53 | 51 (17-87) | 40% | 60% |
| **SARS-CoV-2 negative** | 105 | 47 (12-88) | 43% | 57% |
| **Systemic Lupus Erythematosus (SLE)** | 26 | 36 (26-68) | 35% | 65% |
| **Lyme disease** | 20 | 47 (19-84) | 45% | 55% |
| **CMV** | 4 | 25 (18-31) | 75% | 25% |
| **Hepatitis C** | 20 | 54 (21-89) | 45% | 55% |
| **Syphilis** | 6 | 53 (33-73) | 33% | 67% |
| **Celiac disease** | 26 | 48 (18-75) | 38% | 62% |
| **Rheumatoid arthritis** | 26 | 52 (21-84) | 46% | 54% |
| **Healthy Controls** | 268 | 35 (19-70) | 34% | 66% |
| ***CROSS REACTIVITY CONTROLS*** |  |  |  |  |
| **ANA (Anti-Nuclear Antibodies)** | 79 | 57 (8-90) | 34% | 66% |
| **HBV antibodies** | 18 | 47 (19-82) | 42% | 58% |
| **HCV antibodies** | 14 | 48 (22-62) | 50% | 50% |
| **Influenza A antibodies** | 42 | 45 (20-65) | 27% | 73% |
| **Influenza B antibodies** | 26 | 48 (22-75) | 38% | 62% |
| **Respiratory Syncytial Virus antibodies** | 52 | 51 (18-78) | 38% | 62% |
| **Common Human Coronavirus** | 8 | 45(21-66) | 48% | 52% |
| **Adenovirus** | 4 | 47 (18-71) | 50% | 50% |
| **Coxsackie Virus** | 31 | 52 (36-78) | 38% | 62% |
| **Echovirus** | 28 | 44 (22-61) | 40% | 60% |
| **Poliovirus** | 11 | 50 (18-80) | 48% | 52% |
| **Rhinovirus** | 4 | 48 (24-66) | 50% | 50% |

### Study Approval and Informed Consent Process

The study was conducted under the ethical principles that have their origins in the Declaration of Helsinki. The serum samples used for investigation studies such as disease controls were obtained from third-party specimen providers under individual IRBs. The collection of samples with NP swab positive and negative results for SARS-CoV2 was done under a multi-site central IRB (IRB # 1-1288754-1). Informed consent form and patient questionnaire approved under the central IRB were used at all sites. Remnant de-identified samples that were collected prior to the disease outbreak were under IRB # 1-1098539-1.

### Pillar Plate Assembly

The test plate used by Vibrant America is in a 96-pillar plate format as shown in Figure 1. Briefly, silicon wafers are pre-processed to make a high binding surface capable of immobilizing proteins. The following antigens were included in the panel: *S1 glycoprotein, Receptor binding domain, S2 glycoprotein, Nucleoprotein*. The recombinant antigens were expressed in HEK293 cell lines using full length cDNA coding for the respective antigens fused with a hexa histidine purification tag. Individual wafers were immobilized with each antigen which were then diced into 0.7×80.7 mm^2^ microchips using a stealth dicing process. The diced wafers were picked and placed onto individual carrier tapes using a standard die sorting system. The carrier tapes were loaded onto a high-throughput surface mount technology (SMT) component placement system. The microchips were then picked and placed onto 96-pillar plates with each pillar containing a layout of 4 microchips – one for each antigen being probed.

### DBS Sample Processing

DMPK-type C, Perkin Elmer 226 and Whatman 903 cards suitable for protein-based analysis were used in this study. The card is compatible with automated punchers enabling rapid scaling depending on processing requirements. Briefly, four punches per card 6mm in diameter were made on the DMPK-C cards and collected in 96 well plates. The elution was done in phosphate buffered saline prior to dilution and performance of the assay.

### Assay Protocol

Serum samples were probed using 1:50 dilution and DBS samples using a 1:20 dilution on the pillar plate and reacted for 15 minutes at room temperature. The plate was then washed with Tris-buffered saline with Tween 20 (TBST) (Amresco) buffer 3 × 5 minutes each. The plate was incubated with the secondary antibody (1:2000 dilution of Goat Anti-Human IgG HRP and Goat Anti-Human IgM HRP and Goat Anti-Human IgA HRP individually) for 15 minutes at room temperature. The plates were then washed with TBST buffer followed by washing with DI Water. The plates were finally dried completely before adding chemiluminescent substrate (Clarity Max from Bio-Rad) and scanned for five minutes on a standard Chemiluminescence Imager. For the Enhanced IgM Assay, the serum was pre reacted with Goat anti-human IgG Fc fragment prior to the remaining assay steps to increase the sensitivity of IgM and IgA detection. Assaying is performed on automated liquid handlers enabling a throughput of 100,000 samples per day.

### Data Analysis

The raw chemiluminescent signals for all the probes were extracted from the images using an in-house reporter software. The chemiluminescent signals were converted into intensity plots after quantile normalization, background and spatial correction. The signal threshold was defined for each antigen by calculating the mean +/− SD of the signal intensity for the same antigen among the healthy controls collected prior to the infection outbreak. The raw data was converted into arbitrary chemiluminescent units (CU) based on each individual antigen cut-off for further analysis.

## RESULTS

### Analytical Performance

We evaluated the analytical performance characteristics of the CoV2 Antibody assay for the following parameters: precision (repeatability/reproducibility), analytical specificity and linearity. Precision (repeatability) was determined by having two test operators run a panel of 6 samples, 4 replicates daily over a period of 5 days for a total of 40 data points per sample. Panel consisted of positive control, negative control, positive sample, negative sample, a sample with concentration +20% above cut-off, and a sample with concentration −20% below cut-off. The results are summarized in Table 2.

**Table 2:**
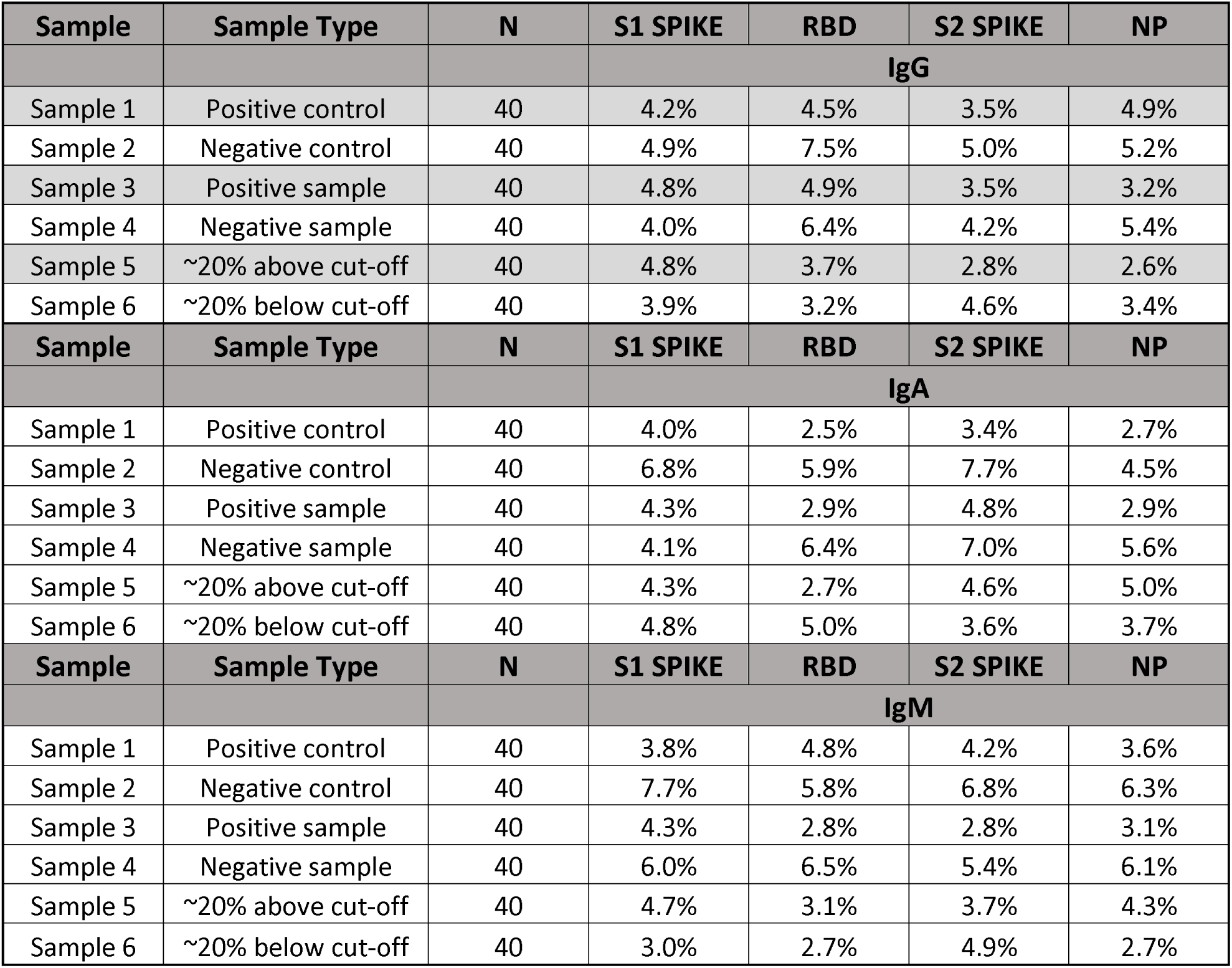
Analytical Reproducibility Summary

| Sample | Sample Type | N | S1 SPIKE | RBD | S2 SPIKE | NP |
| --- | --- | --- | --- | --- | --- | --- |
| IgG |  |  |  |  |  |  |
| Sample 1 | Positive control | 40 | 4.2% | 4.5% | 3.5% | 4.9% |
| Sample 2 | Negative control | 40 | 4.9% | 7.5% | 5.0% | 5.2% |
| Sample 3 | Positive sample | 40 | 4.8% | 4.9% | 3.5% | 3.2% |
| Sample 4 | Negative sample | 40 | 4.0% | 6.4% | 4.2% | 5.4% |
| Sample 5 | ~20% above cut-off | 40 | 4.8% | 3.7% | 2.8% | 2.6% |
| Sample 6 | ~20% below cut-off | 40 | 3.9% | 3.2% | 4.6% | 3.4% |
| Sample | Sample Type | N | S1 SPIKE | RBD | S2 SPIKE | NP |
| IgA |  |  |  |  |  |  |
| Sample 1 | Positive control | 40 | 4.0% | 2.5% | 3.4% | 2.7% |
| Sample 2 | Negative control | 40 | 6.8% | 5.9% | 7.7% | 4.5% |
| Sample 3 | Positive sample | 40 | 4.3% | 2.9% | 4.8% | 2.9% |
| Sample 4 | Negative sample | 40 | 4.1% | 6.4% | 7.0% | 5.6% |
| Sample 5 | ~20% above cut-off | 40 | 4.3% | 2.7% | 4.6% | 5.0% |
| Sample 6 | ~20% below cut-off | 40 | 4.8% | 5.0% | 3.6% | 3.7% |
| Sample | Sample Type | N | S1 SPIKE | RBD | S2 SPIKE | NP |
| IgM |  |  |  |  |  |  |
| Sample 1 | Positive control | 40 | 3.8% | 4.8% | 4.2% | 3.6% |
| Sample 2 | Negative control | 40 | 7.7% | 5.8% | 6.8% | 6.3% |
| Sample 3 | Positive sample | 40 | 4.3% | 2.8% | 2.8% | 3.1% |
| Sample 4 | Negative sample | 40 | 6.0% | 6.5% | 5.4% | 6.1% |
| Sample 5 | ~20% above cut-off | 40 | 4.7% | 3.1% | 3.7% | 4.3% |
| Sample 6 | ~20% below cut-off | 40 | 3.0% | 2.7% | 4.9% | 2.7% |

Furthermore, an interfering substance study was conducted to evaluate the potential interference of specific endogenous and exogenous substances to determine analytical specificity. The interfering substances with levels tested include Bilirubin 40 mg/dl, Cholesterol 100 mg/dl, Triglycerides 1000 mg/dl, Hemoglobin 1000 mg/dl, Rheumatoid Factor (RF) 2000 IU/ml, HAMA 12.5ng/ml, Ribavirin 25mg/dl, Levofloxacin .5mg/dl, Azithromycin .5mg/dl, Ceftriaxone sodium 25mg/dl, Oxymetazoline 1.25mg/dl, Sodium chloride 25mg/dl, EDTA 12.5mg/ml, Acetaminophen 50mg/ml, Ibuprofen 50mg/ml, Budesonide 1.25mg/dl. No interference was observed with any of the substances tested at the stated levels.

Linearity and recovery were tested by diluting positive samples across the assay measuring range for each antigen in six serial dilutions with negative patient sera. Samples were assayed by adding mixtures of positive sample and spiking varying concentration of negative sample to obtain a serial dilution curve. Recoveries for spiked test samples were calculated by comparison to the measured recovery of undiluted results. All values represent the average of three replicates tested. The observed values were evaluated against the calculated theoretical values and a linear regression analysis was performed. R^2^ regression values were greater than 0.98 for all antigens tested. The results are summarized in Table 3.

**Table 3:**
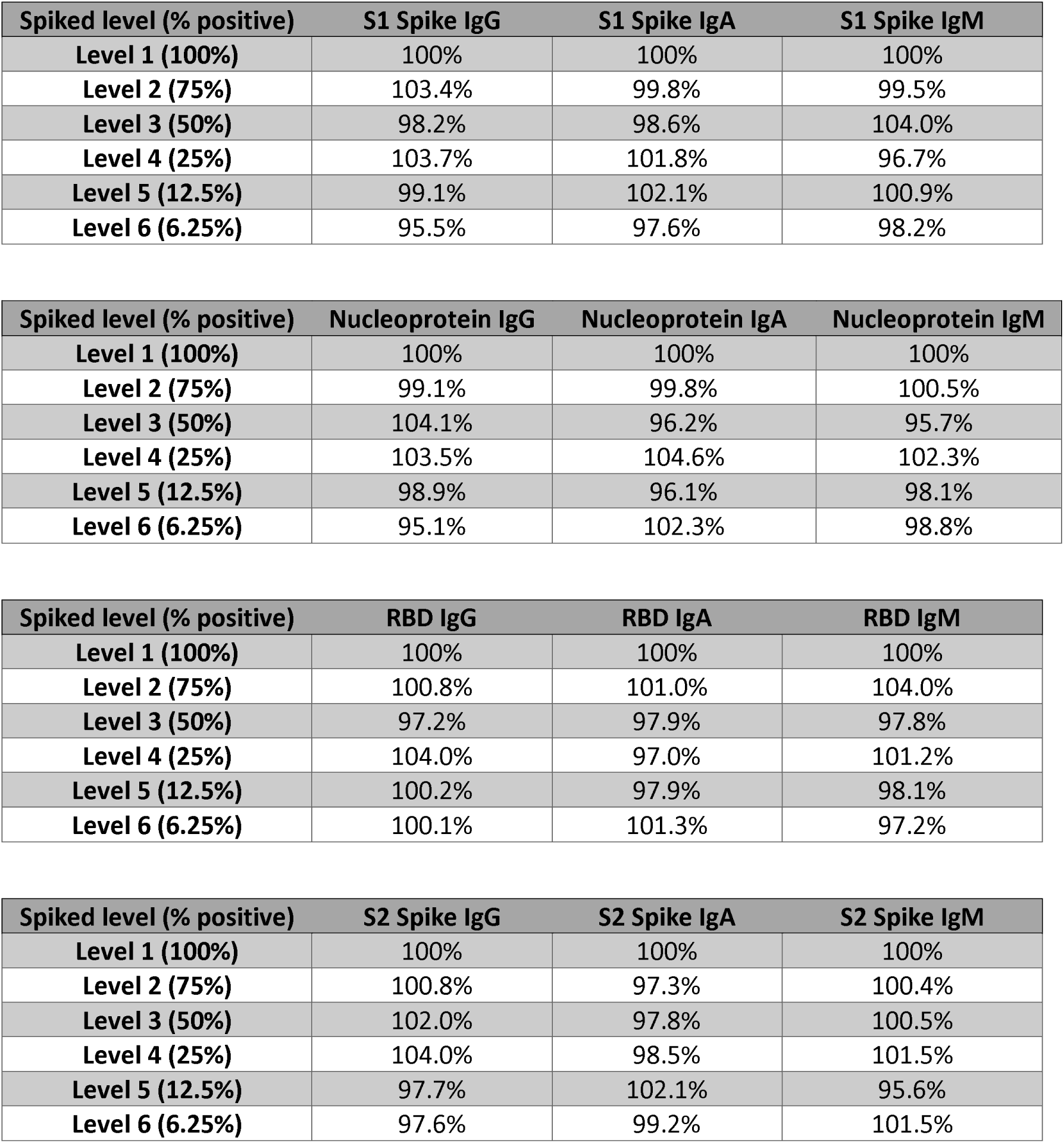
Linearity Studies

### Clinical Sensitivity and Specificity

The clinical study to determine the sensitivity and specificity was done using a panel containing retrospectively collected patient serum samples that have been previously confirmed to be positive by SARS-CoV-2 RT PCR along with healthy controls (samples collected prior to SARS-CoV-2 outbreak across age groups) and other disease controls including autoimmune and infectious diseases(as listed in Table 1). Importantly virus positive samples including for other common coronaviruses, influenza A, B, RSV, echovirus, rhinovirus, adenovirus, poliovirus, coxsackie virus, rhinovirus, hepatitis B and hepatitis C did not show cross reactivity to the four antigens specific to the novel coronavirus. Table 4 summarizes the sensitivity and specificity of the individual antigens across the subclasses of antibodies. The specificity of each antigen is relatively high, up to 99%, but the sensitivity is varied among tested antigens. Three antigens - S2 spike IgG, S1 spike IgM, and S2 spike IgM showed more than 80% of sensitivity to identify the cases. When combining all antigens and antibody subclasses together, the CoV2 antibody assay showed 98.1% sensitivity and 98.6% specificity.

**Table 4:** Clinical Performance Summary

| **Antigen** | **Clinical Sensitivity**<br>**(95% confidence)** | **Clinical Specificity**<br>**(95% confidence)** |
| --- | --- | --- |
| S1 Spike IgG | 75.5% (62.4%-85.1%) | 98.8% (97.4%-99.5%) |
| RBD IgG | 64.2% (50.7%-75.7%) | 99.2% (97.9%-99.7%) |
| S2 Spike IgG | 84.9% (72.9%-92.2%) | 99.4% (98.3%-99.8%) |
| NP IgG | 71.7% (58.4%-82.0%) | 99.0% (97.7%-99.6%) |
| S1 Spike IgA | 56.6% (43.3%-69.1%) | 99.4% (98.3%-99.8%) |
| RBD IgA | 47.2% (34.4%-60.3%) | 99.2% (97.9%-99.7%) |
| S2 Spike IgA | 49.1% (36.1%-62.1%) | 99.6% (98.6%-99.9%) |
| NP IgA | 37.7% (25.9%-51.1%) | 99.2% (97.9%-99.7%) |
| S1 Spike IgM | 81.1% (68.6%-89.4%) | 98.8% (97.4%-99.5%) |
| RBD IgM | 79.3% (66.5%-88.0%) | 99.2% (97.9%-99.7%) |
| S2 Spike IgM | 84.9% (72.9%-92.2%) | 99.6% (98.6%-99.9%) |
| NP IgM | 67.9% (54.5%-78.9%) | 99.2% (97.9%-99.7%) |
| **OVERALL** | **98.1% (90.1%-99.7%)** | **98.6% (97.1%-99.3%)** |

### Matrix Comparison – SST vs DBS

Dried blood spots are a convenient method of collection and processing of samples in pandemic settings. They can be collected in isolation by the affected subject thereby limiting the chances of spreading the infection. Additionally, they do not require any centrifuge for pre-processing and are stable at room temperature for days. Central labs can process these specimens with automated punchers enabling scalable throughput. Specific filter papers for different applications have been developed and validated including their use in measurement of antibodies to viruses. Prospective collection of 1418 samples at various clinical sites of both venipuncture-based SST collection and dried blood spot collection was performed and results summarized in supplementary figure 1 along with 158 clinically paired samples. The reproducibility was greater than 98% across antigens and antibody subtypes.

### CoV2 Antibody positivity in US population

This testing platform with the ability to detect three antibody subclasses – IgM, IgA and IgG to four key antigens is an FDA notified assay. A submission has been made for EUA and testing of these antibodies is currently being performed across the US. More than 7700 individuals have tested from different geographies and their summary data is shown in Table 5. The range of IgA positivity to CoV2 antigens was from 3 to 21%, which was lower than IgM or IgG positivity. The overall positivity rate of any antibody for the tested population was 20.4%.

**Table 5:** Antibody Prevalence in the US Population

| State | Total Samples Tested | Any antibody positive N (%) | IgM positive N (%) | IgA positive N (%) | IgG positive N (%) |
| --- | --- | --- | --- | --- | --- |
| AZ | 1290 | 268 (20.8%) | 187 (14.5%) | 48 (3.7%) | 136 (10.5%) |
| CA | 2518 | 500 (19.9%) | 319 (12.7%) | 105 (4.2%) | 291 (11.6%) |
| CO | 1191 | 257 (21.6%) | 165 (13.9%) | 66 (5.5%) | 159 (13.4%) |
| FL | 236 | 46 (19.5%) | 25 (10.6%) | 14 (5.9%) | 29 (12.3%) |
| GA | 126 | 33 (26.2%) | 21 (16.7%) | 9 (7.1%) | 29 (23.0%) |
| IL | 241 | 37 (15.4%) | 24 (10.0%) | 8 (3.3%) | 26 (10.8%) |
| IN | 127 | 15 (11.8%) | 10 (7.9%) | 3 (2.4%) | 6 (4.7%) |
| KS | 117 | 31 (26.5%) | 12 (10.3%) | 7 (6.0%) | 25 (21.4%) |
| MA | 145 | 44 (30.3%) | 27 (18.6%) | 8 (5.5%) | 26 (17.9%) |
| ME | 25 | 4 (16.0%) | 4 (16.0%) | 0 (0.0%) | 0 (0.0%) |
| MI | 61 | 16 (26.2%) | 10 (16.4%) | 7 (11.5%) | 10 (16.4%) |
| MN | 48 | 7 (14.6%) | 4 (8.3%) | 3 (6.3%) | 4 (8.3%) |
| MO | 161 | 24 (14.9%) | 14 (8.7%) | 5 (3.1%) | 12 (7.5%) |
| NC | 52 | 7 (13.5%) | 7 (13.5%) | 1 (1.9%) | 1 (1.9%) |
| NE | 102 | 19 (18.6%) | 10 (9.8%) | 7 (6.9%) | 14 (13.7%) |
| NJ | 343 | 103 (30.0%) | 70 (20.4%) | 40 (11.7%) | 75 (21.9%) |
| NY | 82 | 29 (35.4%) | 13 (15.9%) | 17 (20.7%) | 27 (32.9%) |
| OH | 47 | 17 (36.2%) | 8 (17.0%) | 5 (10.6%) | 10 (21.3%) |
| OR | 107 | 16 (15.0%) | 8 (7.5%) | 5 (4.7%) | 11 (10.3%) |
| PA | 61 | 5 (8.2%) | 3 (4.9%) | 2 (3.3%) | 3 (4.9%) |
| TX | 633 | 94 (14.8%) | 70 (11.1%) | 24 (3.8%) | 53 (8.4%) |
| WA | 29 | 5 (17.2%) | 2 (6.9%) | 1 (3.4%) | 3 (10.3%) |
| WI | 18 | 3 (16.7%) | 3 (16.7%) | 0 (0.0%) | 0 (0.0%) |
| WY | 13 | 4 (30.8%) | 2 (15.4%) | 3 (23.1%) | 3 (23.1%) |
| Grand Total | 7773 | 1584 (20.4%) | 1018 (13.1%) | 388 (5.0%) | 953 (12.3%) |

## DISCUSSION

Serological assays play a critical role in identifying burden of disease and also to determine exposure and potential immunity against the infectious agent in a population to enable resumption of economic activities. In a pandemic setting, discreet collection of samples and high-volume processing in central laboratories are essential for containment and understanding of disease burden. A dried blood spot-based collection and high-volume testing using a flexible platform to test for antibodies of all subclasses against multiple antigens was validated in this study. The ability to add novel discovered antigens and continuously measuring the antibody levels across IgM, IgA and IgG subclasses enable real-time epidemiology studies in a disease outbreak using this platform.

Our data shows 100% seroconversion in individuals infected with SARS-CoV-2. Median seroconversion time was seven days from the day of microbial swab confirmation. Overall clinical sensitivity was 98.1% with a specificity of 98.6% while S1 and S2 Spike proteins were the two most sensitive markers observed. Currently diagnosis of the SARS-CoV2 infection is predominantly based on NP swab-based confirmation of microbial DNA while serology adds value in supplemental testing [11]. Positivity is dependent on sufficient amount of sample present at the sampling site that can be amplified and detected using standard PCR. The time of sampling is also a crucial factor since viral replication and load increases in specific window periods and sampling outside of this may lead to false negatives. Poor techniques in sample collection could also lead to false negatives.

When it comes to serological tests the main limitations that make them suboptimal tools for diagnosing those who are sick is that it takes time for the development of antibodies after infection. Thus, they may produce a false negative result in individuals who are acutely infected. On the other hand, they can be useful to indicate exposure in individuals who are asymptomatic or were symptomatic but PCR negative or were never tested. Nevertheless, it is important to note that the serological testing should not be used as the sole basis for diagnosis of an acute COVID19 infection.

Technology developments truly enable testing for multiple antibodies from small volumes of collected blood. The potential convenience and cost savings that it would bring to laboratory testing cannot be understated. Remote sampling such as with DBS cards, along with telemedicine would help providers to order tests not only in pandemic settings but also can be a great enabler in bringing healthcare costs down. Immediate application of such a platform to do multiplex testing would be in the monitoring of exposure to upper respiratory tract infections.

A subset of the NP swab positive individuals are being monitored for antibody levels and symptoms at regular intervals from the time they tested positive. Initial results show that the seroconversion from IgM to IgG which is marked by a reduction in IgM titer might be co-related with recovery since it is accompanied by a negative result in the subsequent NP swab test and also improvement in symptoms. This could possibly evolve into the criteria for confirmation of recovery from infection which would enable return to work of affected individual.

In summary, we have developed and validated a highly accurate assay that enables the identification of individuals who have been infected with SARS CoV2. The assay meets the rigorous requirements of high sensitivity and specificity, scalability, high fidelity and reliability that will be critical to the control of the current pandemic. The utility of the self-collection method aligns with the dual needs of safe convenient testing with continued isolation for patients that may either be infectious or those who are sheltering at home to avoid infection exposure. If convalescent antibodies are demonstrated to be protective from future infection, then this testing could enable an easing of restrictions both at the individual and community level.

## Data Availability

All data referred to in the manuscript are available and are part of the manuscript

## ACKNOWLEGMENTS

The authors thank the FDA and the staff of CDC for early guidance on validation requirements. We thank the healthcare providers and the study participants who made this research possible. Special thanks to Inna Yaskin, Jaclyn McCoy, Tina Wilson, Roanne Houck and Gunnison Valley Hospital for coordinating research efforts. Research reported in this publication was supported by the Vibrant America LLC. The content is solely the responsibility of the authors and does not necessarily represent the official views of the CDC or FDA. We also would like to thank Miaomiao Sun and Ashish Gopalakrishnan for generating the Figure 1 illustration.

## Author Contributions

Conceived and designed the experiments: HKK VJ KK. Performed the experiments: TW KK. Analyzed the data: KK KER VJ TW RSC. Contributed reagents/materials/analysis tools: TW KB IY GVH. Wrote the paper: HKK VJ. Revised the manuscript critically for important intellectual content: JJR HKK AJR RSC JAM

---

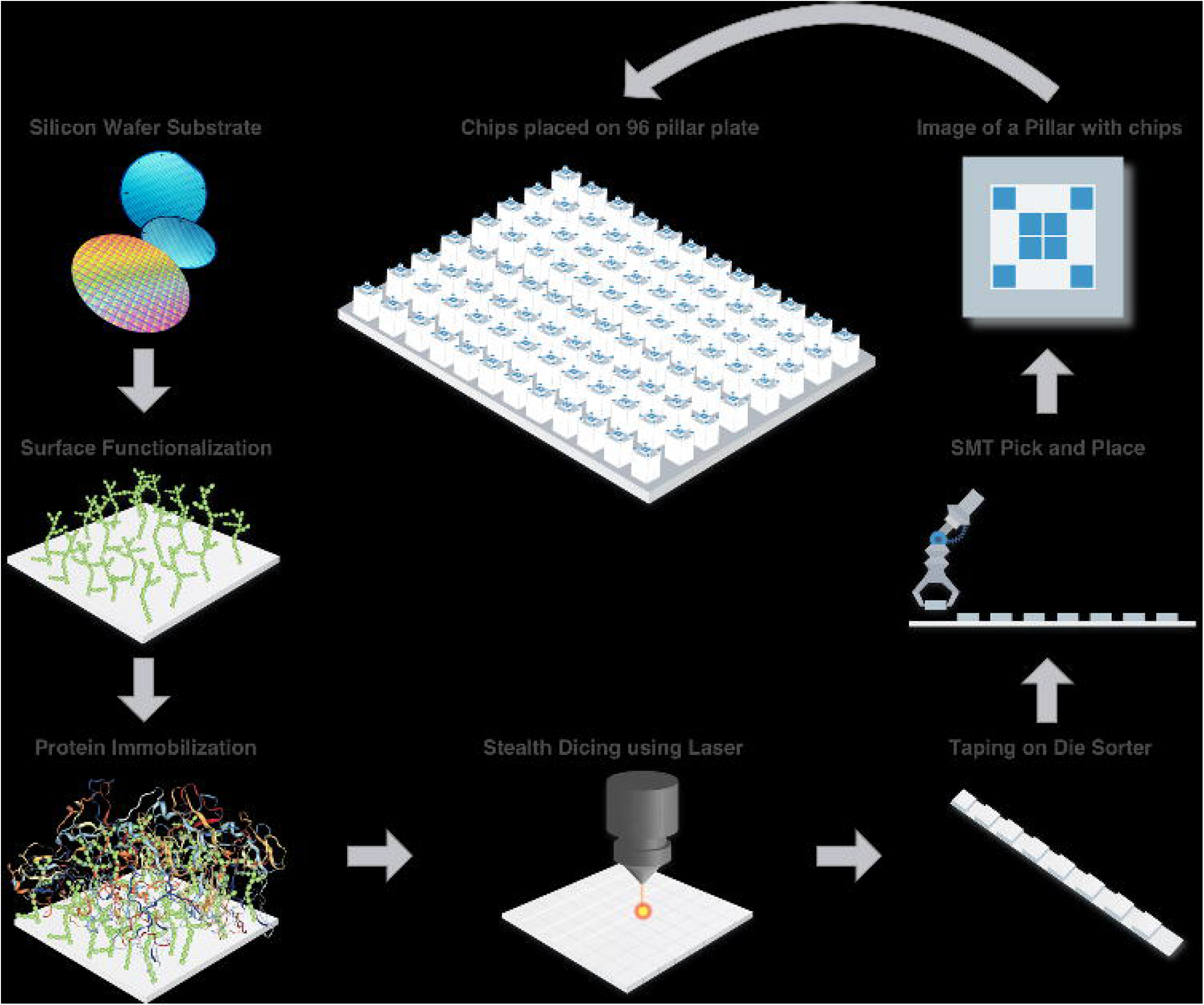

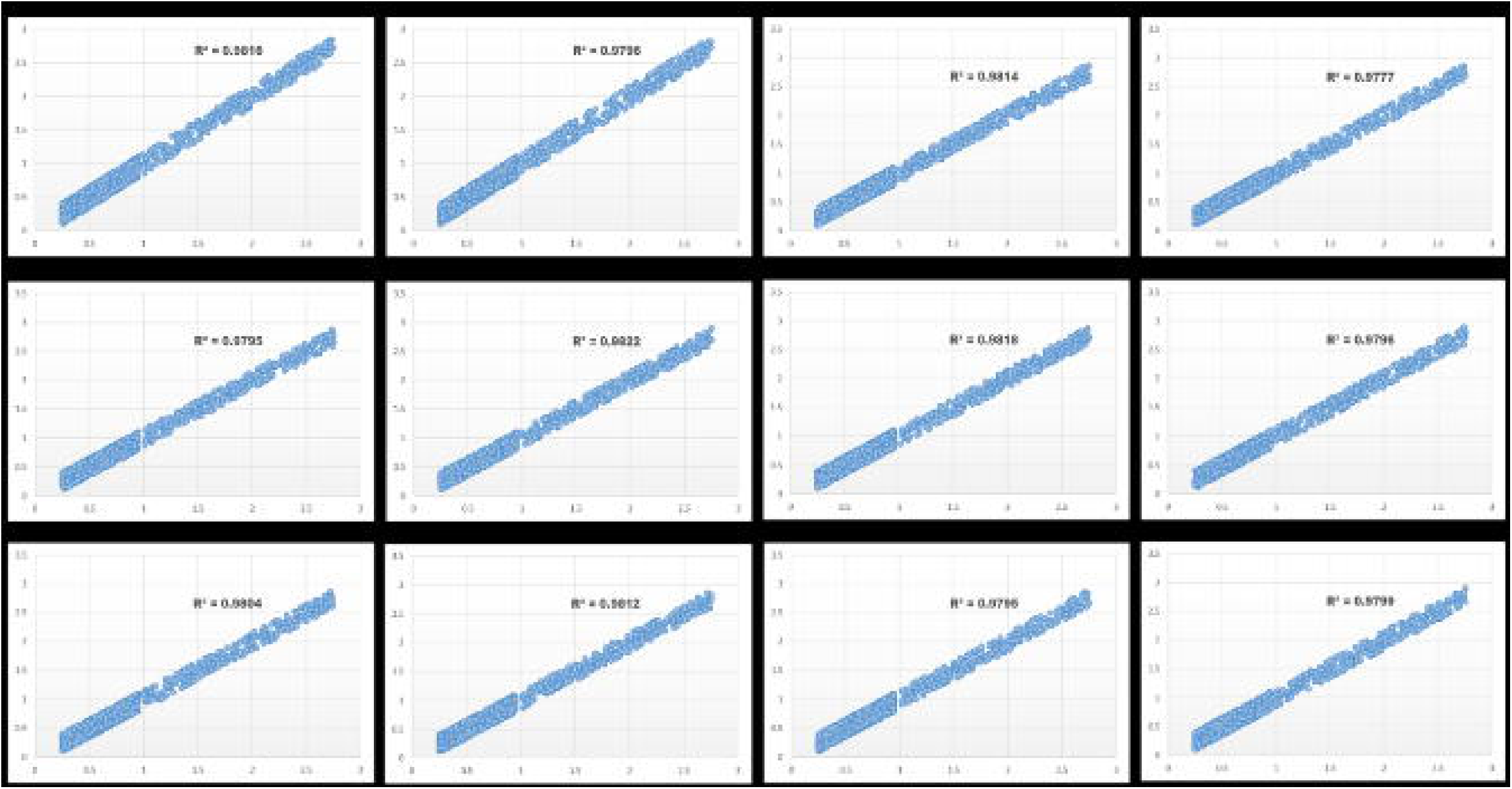

